# Effectiveness of Carotid Sinus Massage in Modified Trendelenburg Position for Rapid and Lasting Relief of Acute Headache Episodes: A Pilot Study

**DOI:** 10.1101/2025.02.11.25321880

**Authors:** Pablo Anaya, Heberto Suarez-Roca

## Abstract

**Introduction:** Headaches significantly impair quality of life, and primary headaches are among the most common neurological complaints. Stimulating carotid baroreceptors via carotid sinus massage (CSM) combined with modified Trendelenburg positioning may modulate autonomic activity and alleviate acute headache symptoms. Thus, this study evaluated the effectiveness and safety of this combined approach (CSM+T).

**Methods:** Seventeen patients (14 females; ages 16–64) with tension-type headaches (6), chronic migraines (10), or mixed-headaches (1) and various comorbidities received up to three 15-second CSM+T applications at one-minute intervals. Pain was assessed using a 10-point visual analog scale (VAS), while heart rate (HR), blood pressure (BP), and oxygen saturation (SpO_2_) were recorded before and after the intervention. A 24-hour follow-up evaluated headache recurrence and adverse effects.

**Results:** Sixteen patients experienced significant pain reduction (median decrease: 9 VAS points; p < 0.0001) without complications. Pain relief persisted 24 hours without the need for additional medication or adverse effects. HR, BP, and SpO_2_ decreased following CSM+T. A negative correlation was observed between HR reduction and pain relief. Among responders, migraine patients had a smaller mean HR decrease than tension-type headache patients, yet both groups achieved similar median pain relief. These results suggest that a more pronounced cardiovagal response does not necessarily confer greater analgesia, implying additional central or multifactorial mechanisms.

**Conclusion:** CSM+T appears to be a safe and effective non-pharmacological intervention for rapid headache relief in a heterogeneous patient population. Larger, controlled trials are warranted to confirm these findings and refine clinical protocols.

## Introduction

Headaches significantly impair quality of life and productivity, and acute episodes contribute substantially to emergency department visits (Filler et al., 2019). Tension-type headaches and migraines are particularly common and are typically managed with pharmacological treatments that risk medication overuse and adverse effects (Chaibi & Russell, 2014). Consequently, effective non-pharmacological interventions are urgently needed.

Recent evidence indicates that blood pressure regulation and baroreceptor function play a crucial role in headache pathophysiology. Higher pulse pressure correlates with reduced risk and prevalence of headaches (Tronvik et al., 2008), while lower baroreflex sensitivity—a measure of how effectively blood pressure changes modulate heart rate—has been observed in affected patients (Virtanen et al., 2004). Moreover, dysregulated baroreflex function, whether due to increased or decreased sensitivity, has been documented in both migraine (Nilsen et al., 2009; Sanya et al., 2005) and tension-type headache patients (Khurana, 2006). Given that baroreceptors in the carotid sinus and aortic arch are pivotal for reflex control of heart rate and blood pressure (Suarez-Roca et al., 2021), these findings underscore their therapeutic potential.

Building on this insight, carotid-based interventions have emerged as promising acute treatments for migraine. For instance, cervical transcutaneous vagus nerve stimulation provides pain relief in approximately 20% of acute migraine cases (Tassorelli et al., 2018). Similarly, applying sliding digital pressure along the common carotid artery—thereby stimulating the carotid sinus—has demonstrated rapid, clinically meaningful pain relief (Tavakoli et al., 2020). However, the duration of relief, the accompanying hemodynamic changes, and the technique’s effectiveness for tension-type headaches remain unclear.

Carotid sinus massage (CSM), traditionally used to diagnose carotid sinus syndrome and treat supraventricular tachycardia (Pasquier et al., 2017), also stimulates baroreceptors that modulate autonomic activity, reduce pain perception, and inhibit muscular contraction (Suarez-Roca et al., 2021), potentially alleviating headache symptoms.

Additionally, the modified Trendelenburg position—where the patient lies supine with a passive 10–30° leg elevation while keeping the head level—temporarily increases mean arterial pressure by enhancing venous return and cardiac output (Ostrow et al., 1994; Terai et al., 1995). This position has been used to improve the efficacy of the Valsalva maneuver in terminating supraventricular tachycardia (Appelboam et al., 2015).

Despite these advances, there remains a need for a more robust and clinically applicable protocol that can provide rapid, sustained headache relief without pharmacological intervention across heterogeneous populations of patients. This pilot study aims to evaluate the effectiveness and safety of combining two well-established techniques—CSM applied in the modified Trendelenburg position—as an acute intervention for migraine and tension-type headaches.

## Methods

### Patient Selection

Seventeen patients (14 females [79%] and 3 males [21%], aged 16– 64 years) presenting with tension-type or mixed headaches at Pedro T. Orellana Hospital in Trenque Lauquen, Argentina, were enrolled. Inclusion criteria were: age ≥16 years, a diagnosis of tension-type headache or migraine per the International Classification of Headache Disorders (ICHD-3), and absence of contraindications for carotid sinus massage (e.g., carotid bruits, recent myocardial infarction, stroke, significant carotid artery stenosis) or for the Trendelenburg position (e.g., increased intracranial pressure, lower limb ischemia, respiratory distress, congestive heart failure), as well as no history of syncope or arrhythmias. The institutional review board approved the study, and all patients provided informed consent (ClinicalTrials.gov Identifier: NCT06745648; C-SMART study).

### Procedure

Patients were placed in the modified Trendelenburg position—supine with legs passively elevated 10°–30°, while continuous monitoring of heart rate, blood pressure, and oxygen saturation (SpO_2_) was performed using a multiparameter monitor (Mindray uMEC12, Shenzhen, China) before, during, and after the procedure. The carotid artery was then identified by palpation, located just below the mandibular angle, above the thyroid cartilage, and medial to the sternocleidomastoid muscle. Gentle, steady circular pressure was applied using the fingertips for 15 seconds without occluding the artery. If sufficient pain relief was not achieved, the massage was repeated after a one-minute interval, up to three times. The right carotid sinus was chosen because its mechanical activation has been shown to slow the heart rate more effectively than the left (Tafil-Klawe et al., 1990).

### Assessment

Pain intensity was evaluated using a 10-point visual analog scale (VAS) at baseline and after each CSM. Hemodynamic parameters—including heart rate, blood pressure, and SpO_2_—were recorded before the first CSM and after the final CSM. A telephone follow-up was conducted 24 hours post-procedure to assess pain recurrence and any delayed adverse effects.

### Statistical Analysis

Changes in VAS scores, heart rate, systolic and diastolic blood pressure, and SpO_2_ were analyzed using the Wilcoxon matched-pairs signed-rank test, appropriate for non-parametric paired data. Linear correlation analysis examined the relationship between VAS pain scores and hemodynamic variables. Statistical significance was set at p < 0.05, and all analyses were performed using SPSS version 29 (IBM Corp., Armonk, NY, USA).

## Results

The cohort exhibited diverse comorbidities, including metabolic disorders (e.g., overweight, type 2 diabetes, insulin resistance), endocrine issues (hypothyroidism), respiratory conditions (mild intermittent asthma, allergic rhinitis), psychological factors (untreated anxiety, smoking), and others (osteoarthritis, endometriosis, dyspepsia), reflecting the population’s heterogeneity. Six patients had tension-type headaches, ten had migraines, and one presented with both. Headache characteristics ranged from tightness or pressure to pulsating pain with photophobia or phonophobia, and locations varied across frontal-temporal, temporal-occipital, occipital, and bilateral regions.

CSM under the modified Trendelenburg position produced complete pain relief in 9 patients (71%) and partial relief (final VAS scores of 1–2) in 4 patients (24%). Patients with partial relief opted against additional analgesic treatment. At the 24-hour follow-up, 16 patients remained pain-free without needing further medication or experiencing delayed adverse effects, while one non-responder required intravenous analgesics after multiple attempts.

In most patients, VAS pain scores dropped markedly from pre-intervention values of 6–10 to 0 after two or three applications **(Table 1)**. One patient maintained a VAS score of 10 despite three attempts but tolerated the procedure without complications. The heart rate decreased in all cases, with reductions ranging from 2 to 24 beats per minute. Both systolic and diastolic blood pressures declined by up to 30 mm Hg, while SpO_2_ decreased by up to 4% yet remained within clinically acceptable ranges. Overall, the intervention was well tolerated without complications.

**Table 1:** VAS Scores and Hemodynamic Parameters Before and After CSM by Case.

| Case | VAS<br>before<br>CSM+T | VAS<br>after 1<sup>st</sup><br>CSM+T | VAS<br>after 2<sup>nd</sup><br>CSM+T | VAS<br>after 3<sup>rd</sup><br>CSM+T | HR<br>before<br>CSM+T | HR<br>after<br>CSM+T | BP<br>before<br>CSM+T | BP<br>after<br>CSM+T | SpO<sub>2</sub><br>before<br>CSM+T | SpO<sub>2</sub><br>after<br>CSM+T |
| --- | --- | --- | --- | --- | --- | --- | --- | --- | --- | --- |
| 1 | 9/10 | 6/10 | 2/10 | N/A | 74 | 65 | 90/40 | 90/40 | 95 | 93 |
| 2 | 9/10 | 6/10 | 2/10 | N/A | 94 | 70 | 100/60 | 100/60 | 98 | 96 |
| 3 | 6/10 | 1/10 | N/A | N/A | 72 | 62 | 140/80 | 120/70 | 96 | 93 |
| 4 | 9/10 | 8/10 | 6/10 | 0/10 | 99 | 84 | 130/80 | 100/70 | 99 | 95 |
| 5 | 9/10 | 7/10 | 4/10 | 0/10 | 82 | 70 | 160/100 | 130/70 | 99 | 98 |
| 6 | 7/10 | 5/10 | 5/10 | 0/10 | 94 | 81 | 150/90 | 130/80 | 99 | 98 |
| 7 | 9/10 | 5/10 | 2/10 | 0/10 | 68 | 64 | 110/80 | 100/70 | 97 | 95 |
| 8 | 10/10 | 7/10 | 4/10 | 0/10 | 78 | 70 | 130/80 | 110/80 | 97 | 96 |
| 9 | 10/10 | 6/10 | 4/10 | 0/10 | 78 | 70 | 110/70 | 110/60 | 98 | 96 |
| 10 | 9/10 | 6/10 | 3/10 | 0/10 | 68 | 58 | 120/70 | 110/60 | 97 | 95 |
| 11 | 10/10 | 6/10 | 4/10 | 0/10 | 68 | 60 | 110/70 | 110/60 | 98 | 96 |
| 12 | 10/10 | 10/10 | 10/10 | 10/10 | 78 | 70 | 110/70 | 110/60 | 98 | 96 |
| 13 | 9/10 | 5/10 | 3/10 | 0/10 | 82 | 74 | 120/70 | 110/60 | 96 | 95 |
| 14 | 8/10 | 6/10 | 4/10 | 1/10 | 96 | 78 | 140/80 | 120/60 | 99 | 99 |
| 15 | 10/10 | 8/10 | 5/10 | 0/10 | 82 | 80 | 120/70 | 110/60 | 98 | 95 |
| 16 | 9/10 | 6/10 | 3/10 | 0/10 | 92 | 86 | 110/80 | 100/80 | 95 | 93 |
| 17 | 10/10 | 5/10 | 2/10 | 0/10 | 90 | 82 | 150/90 | 140/80 | 98 | 96 |

**Note:** HR (heart rate in bpm), BP (systolic/diastolic blood pressure in mmHg), and SpO<sub>2</sub> (% blood oxygen saturation) were measured before the first and after the final CSM+T (carotid sinus massage in the Trendelenburg position). "N/A" indicates that subsequent CSM+T procedures were not performed.

Quantitatively, pain VAS scores decreased by an average of 8.1 units (95% CI: 6.8–9.4; median: 9). Additionally, there were average reductions in heart rate of 10.1 bpm (95% CI: 7.4 to 12.8), systolic blood pressure by 11.8 mm Hg (95% CI: 6.5 to 17.0), diastolic blood pressure by 9.4 mm Hg (95% CI: 5.6 to 13.3), and SpO_2_ by 1.9 % (95% CI: 1.4 to 2.4). The Wilcoxon matched-pairs signed-rank test revealed that reductions in pain intensity VAS scores (P < 0.0001), heart rate (P < 0.0001), systolic blood pressure (P = 0.0005), diastolic blood pressure (P = 0.0002), and SpO_2_ (P < 0.0001) were statistically significant. The pulse pressure change was not significant (−2.4 mm Hg; 95% CI: −7.0 to 2.3; P = 0.3438) **(Figure 1)**.

**Figure 1.**
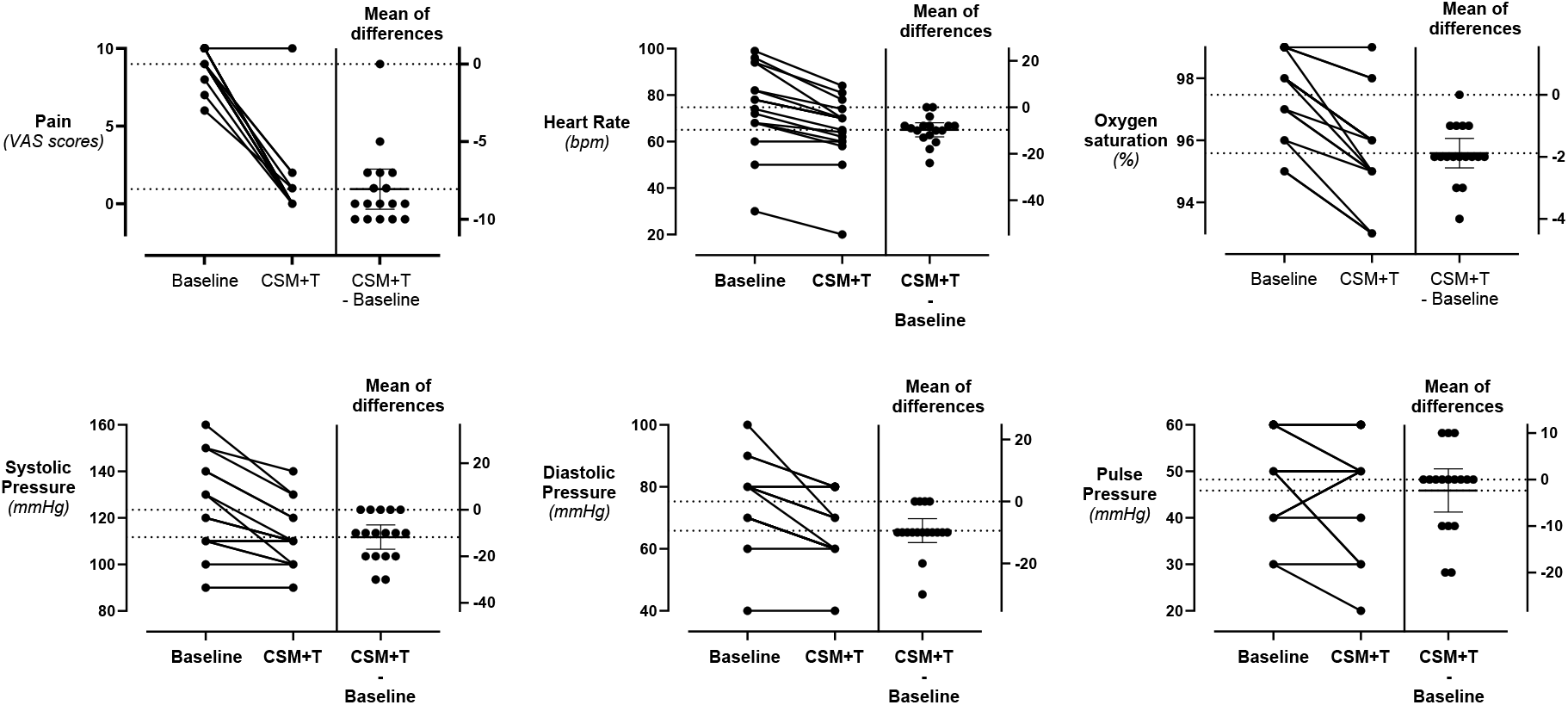
Post-intervention changes in pain, hemodynamics, and SpO_2_. Each subplot displays baseline, post-intervention measures (CSM+T), and the mean differences of each patient’s VAS scores, hemodynamic variables, and SpO_2_. Except for pulse pressure, all means of difference were statistically significant (p < 0.05, Wilcoxon matched-pairs signed-rank test, N=17). Due to overlapping values, some individual changes are not visually distinguishable.

Among intervention responders, pain relief exhibited a negative linear correlation with the post-intervention drop in heart rate (r = - 0.595, p = 0.0168). In contrast, changes in blood pressure and SpO_2_ did not significantly correlate with pain relief **(Figure 2)**.

**Figure 2.**
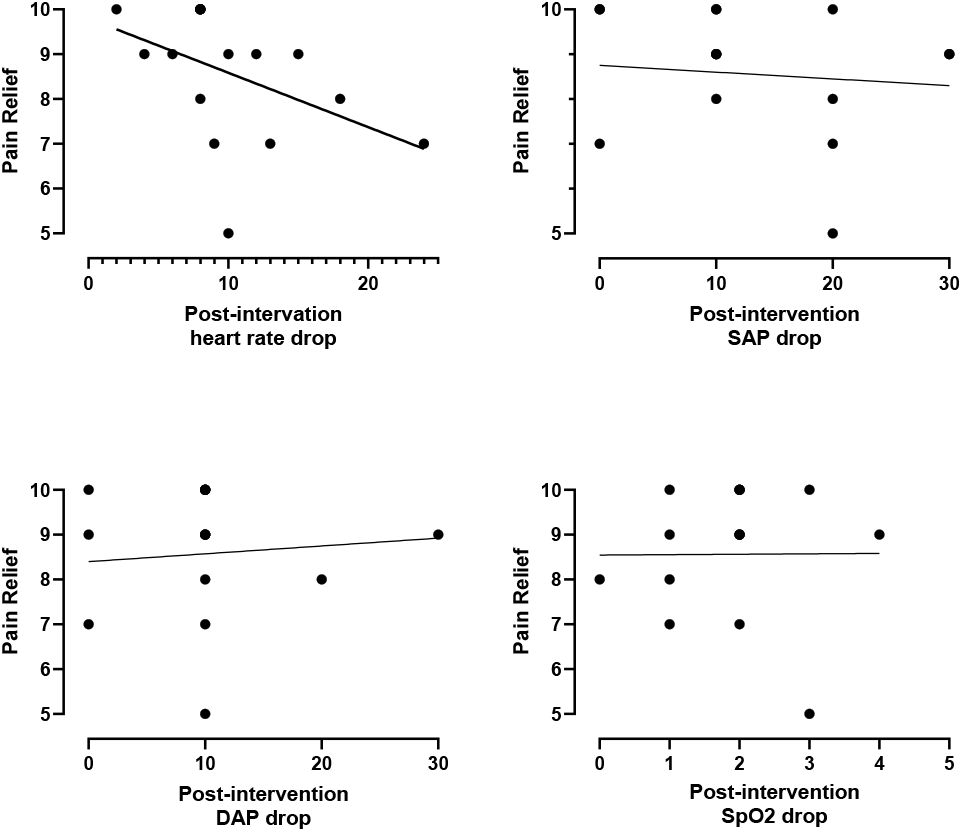
Correlation between pain relief and post-intervention changes in heart rate, blood pressure, and SpO_2_ in patients responding to CSM+T. Pain relief was defined as the difference between baseline and post-intervention scores. The upper left panel depicts a significant negative linear correlation between pain relief and post-intervention drop in heart rate (r = - 0.595, p = 0.0168). The other panels display the lack of a significant correlation between pain relief and changes in blood pressure and SpO_2_. Due to overlapping values, some individual changes are not visually distinguishable.

## Discussion

### Summary of Findings

This pilot study demonstrates that carotid sinus massage in the modified Trendelenburg position provided rapid and significant headache relief in 16 of 17 patients, with a median VAS score reduction of 9 points. The high success rate across diverse ages, sexes, headache types, and comorbidities suggests that this combined intervention may be effective in varied clinical populations. One patient did not experience relief, possibly due to factors such as overweight status (Zhuang et al., 2024), younger age, lower pulse pressure (Tronvik et al., 2008), or prolonged self-medication with analgesics – factors known to be associated with more refractory headache presentations. These results underscore the likely influence of individual variability in treatment response.

### Comparison With Other Treatments

Pharmacological interventions such as triptans and NSAIDs typically relieve migraines in 30–60% of patients within two hours (Ferrari et al., 2001; Moore et al., 2014) but are limited by overuse and adverse effects. Non-pharmacological approaches, including cervical transcutaneous vagus nerve stimulation, have shown only modest and time-dependent benefits – approximately 21% relief at two hours in early studies (Goadsby et al., 2014) and benefits persisting only up to 60 minutes in later trials (Tassorelli et al., 2018). Manual therapies like physiotherapy and relaxation exercises, though effective over time in reducing headache frequency and severity (Chaibi & Russell, 2014; Corum et al., 2021), require prolonged administration and are impractical for acute attacks. In contrast, digital compression techniques targeting extracranial and carotid arteries have provided rapid relief in 68–90% of patients (Hmaidan & Cianchetti, 2006; Tavakoli et al., 2020), although the duration of benefit remains uncertain. Our findings suggest that combining CSM with the modified Trendelenburg position may enhance and extend the therapeutic benefits of manual interventions—a concept supported by similar benefits observed with semi-recumbent positioning during vagal maneuvers for supraventricular tachycardia (Appelboam et al., 2015).

### Mechanism of Action

The reductions in pain intensity, heart rate, and blood pressure, alongside minimal changes in oxygen saturation, support a mechanism involving baroreceptor activation. This activation likely increases parasympathetic tone and decreases sympathetic activity (Suarez-Roca et al., 2019). Additionally, the modified Trendelenburg position may augment venous return and cardiac output (Terai et al., 1995), thereby enhancing carotid sinus stretch and intensifying baroreceptor stimulation. The resultant effects – analgesia, vasodilation, and muscle relaxation – may together contribute to headache relief (Suarez-Roca et al., 2021).

A notable observation was the negative correlation between the magnitude of heart rate reduction and pain relief. Patients with a more pronounced heart rate drop, indicative of a robust baroreflex, paradoxically experienced less analgesic benefit. Consistent with previous reports of altered baroreflex responses in migraine patients (Sanya et al., 2005), our data revealed that migraine patients exhibited a lower mean heart rate decrease (8.4 ± 4.2 bpm) compared to tension-type headache patients (13 ± 4.2 bpm), yet both groups achieved similar median pain relief (VAS scores: 10 vs. 9.5). This finding suggests that stronger cardiovagal activation does not necessarily translate into enhanced analgesia, underscoring the multifactorial nature of headache relief.

### Safety and Tolerability

No adverse events were reported, and the slight reduction in oxygen saturation – likely reflecting enhanced parasympathetic drive and minor decreases in respiratory effort (Zhao et al., 1992) – was clinically insignificant. The modified Trendelenburg position may have contributed to hemodynamic stability during CSM, and proper patient selection and technique appear to mitigate potential risks, in line with previous reports on the safety of CSM (Pasquier et al., 2017; Ungar et al., 2016).

### Limitations

Despite these promising results, several limitations must be acknowledged. The single-center, unblinded design without a sham comparator limits the generalizability of our findings and raises the possibility of placebo effects or spontaneous headache fluctuations. The small sample size, reliance on subjective VAS scores, and short 24-hour follow-up further constrain our conclusions. Future randomized, controlled trials with longer follow-ups, objective outcome measures, and formal baroreflex assessments are needed to confirm these preliminary findings.

## Conclusion

In this pilot study, CSM combined with the modified Trendelenburg position provided rapid and significant relief of tension-type headaches and migraines without adverse effects. Although preliminary, these findings suggest that this combined technique could serve as a non-pharmacological, accessible, and cost-effective option for immediate headache relief in emergency settings. Further research is required to confirm its efficacy, elucidate its underlying mechanisms, and establish standardized clinical protocols.

## Acknowledgments

The authors thank the patients for their participation and consent to publish these cases. We also acknowledge Dr. Jesus Estevez for his valuable comments on earlier versions of the manuscript.

## Data Availability

All data produced in the present work are contained in the manuscript.

## References

Appelboam, A., Reuben, A., Mann, C., Gagg, J., Ewings, P., Barton, A., Lobban, T., Dayer, M., Vickery, J., & Benger, J. (2015). Postural modification to the standard Valsalva manoeuvre for emergency treatment of supraventricular tachycardias (REVERT): a randomised controlled trial. Lancet, 386(10005), 1747–1753. 10.1016/s0140-6736(15)61485-4

Chaibi, A., & Russell, M. B. (2014). Manual therapies for primary chronic headaches: a systematic review of randomized controlled trials. J Headache Pain, 15(1), 67. 10.1186/1129-2377-15-67

Corum, M., Aydin, T., Medin Ceylan, C., & Kesiktas, F. N. (2021). The comparative effects of spinal manipulation, myofascial release and exercise in tension-type headache patients with neck pain: A randomized controlled trial. Complement Ther Clin Pract, 43, 101319. 10.1016/j.ctcp.2021.101319

Ferrari, M. D., Roon, K. I., Lipton, R. B., & Goadsby, P. J. (2001). Oral triptans (serotonin 5-HT(1B/1D) agonists) in acute migraine treatment: a meta-analysis of 53 trials. Lancet, 358(9294), 1668–1675. 10.1016/s0140-6736(01)06711-3

Filler, L., Akhter, M., & Nimlos, P. (2019). Evaluation and Management of the Emergency Department Headache. Semin Neurol, 39(1), 20–26. 10.1055/s-0038-1677023

Goadsby, P. J., Grosberg, B. M., Mauskop, A., Cady, R., & Simmons, K. A. (2014). Effect of noninvasive vagus nerve stimulation on acute migraine: an open-label pilot study. Cephalalgia, 34(12), 986–993. 10.1177/0333102414524494

Hmaidan, Y., & Cianchetti, C. (2006). Effectiveness of a prolonged compression of scalp arteries on migraine attacks. J Neurol, 253(6), 811–812. 10.1007/s00415-006-0085-3

Khurana, R. K. (2006). Headache in Patients with Baroreflex Failure. Headache: The Journal of Head and Face Pain, 46(7), 1207-1209. 10.1111/j.1526-4610.2006.00514_3.x

Moore, R. A., Derry, S., Wiffen, P. J., Straube, S., & Bendtsen, L. (2014). Evidence for efficacy of acute treatment of episodic tension-type headache: methodological critique of randomised trials for oral treatments. Pain, 155(11), 2220–2228. 10.1016/j.pain.2014.08.009

Nilsen, K. B., Tronvik, E., Sand, T., Gravdahl, G. B., & Stovner, L. J. (2009). Increased baroreflex sensitivity and heart rate variability in migraine patients. Acta Neurol Scand, 120(6), 418–423. 10.1111/j.1600-0404.2009.01173.x

Ostrow, C. L., Hupp, E., & Topjian, D. (1994). The effect of Trendelenburg and modified trendelenburg positions on cardiac output, blood pressure, and oxygenation: a preliminary study. Am J Crit Care, 3(5), 382–386. 10.4037/ajcc1994.3.5.382

Pasquier, M., Clair, M., Pruvot, E., Hugli, O., & Carron, P. N. (2017). Carotid Sinus Massage. N Engl J Med, 377(15), e21. 10.1056/NEJMvcm1313338

Sanya, E. O., Brown, C. M., von Wilmowsky, C., Neundörfer, B., & Hilz, M. J. (2005). Impairment of parasympathetic baroreflex responses in migraine patients. Acta Neurol Scand, 111(2), 102–107. 10.1111/j.1600-0404.2004.00358.x

Suarez-Roca, H., Klinger, R. Y., Podgoreanu, M. V., Ji, R. R., Sigurdsson, M. I., Waldron, N., Mathew, J. P., & Maixner, W. (2019). Contribution of Baroreceptor Function to Pain Perception and Perioperative Outcomes. Anesthesiology, 130(4), 634–650. 10.1097/aln.0000000000002510

Suarez-Roca, H., Mamoun, N., Sigurdson, M. I., & Maixner, W. (2021). Baroreceptor Modulation of the Cardiovascular System, Pain, Consciousness, and Cognition. Compr Physiol, 11(2), 1373–1423. 10.1002/cphy.c190038

Tafil-Klawe, M., Raschke, F., & Hildebrandt, G. (1990). Functional asymmetry in carotid sinus cardiac reflexes in humans. Eur J Appl Physiol Occup Physiol, 60(5), 402–405.

Tassorelli, C., Grazzi, L., de Tommaso, M., Pierangeli, G., Martelletti, P., Rainero, I., Dorlas, S., Geppetti, P., Ambrosini, A., Sarchielli, P., Liebler, E., & Barbanti, P. (2018). Noninvasive vagus nerve stimulation as acute therapy for migraine: The randomized PRESTO study. Neurology, 91(4), e364–e373. 10.1212/wnl.0000000000005857

Tavakoli, F., Tafakhori, A., Zebardast, J., & Khan, Z. H. (2020). A new technique employing direct tactile pressure on the common carotid artery to relieve acute episode attack of migraine headache: a single-arm interventional study. Frontiers in Emergency Medicine, 5(1), 7. 10.22114/ajem.v0i0.267

Terai, C., Anada, H., Matsushima, S., Shimizu, S., & Okada, Y. (1995). Effects of mild Trendelenburg on central hemodynamics and internal jugular vein velocity, cross-sectional area, and flow. Am J Emerg Med, 13(3), 255–258. 10.1016/0735-6757(95)90194-9

Tronvik, E., Stovner, L. J., Hagen, K., Holmen, J., & Zwart, J. A. (2008). High pulse pressure protects against headache: prospective and cross-sectional data (HUNT study). Neurology, 70(16), 1329–1336. 10.1212/01.wnl.0000309222.79376.57

Ungar, A., Rivasi, G., Rafanelli, M., Toffanello, G., Mussi, C., Ceccofiglio, A., McDonagh, R., Drumm, B., Marchionni, N., Alboni, P., & Kenny, R. A. (2016). Safety and tolerability of Tilt Testing and Carotid Sinus Massage in the octogenarians. Age Ageing, 45(2), 242–248. 10.1093/ageing/afw004

Virtanen, R., Jula, A., Huikuri, H., Kuusela, T., Helenius, H., Ylitalo, A., Voipio-Pulkki, L. M., Kauma, H., Kesäniemi, Y. A., & Airaksinen, J. (2004). Increased pulse pressure is associated with reduced baroreflex sensitivity. J Hum Hypertens, 18(4), 247–252. 10.1038/sj.jhh.1001661

Zhao, J. M., Sand, T., & Sjaastad, O. (1992). Cluster headache: oxygen saturation and end-tidal CO2 during and without attack. Headache, 32(3), 126–131. 10.1111/j.1526-4610.1992.hed3203126.x

Zhuang, C., Mao, J., Ye, H., He, J., Hu, Y., Hu, H., & Zheng, Y. (2024). Association between severe headache or migraine and lipid accumulation product and visceral adiposity index in adults: a cross-sectional study from NHANES. Lipids Health Dis, 23(1), 307. 10.1186/s12944-024-02303-w

